# CausalFund: Causality-Inspired Domain Generalization in Retinal Fundus Imaging for Low-Resource Screening

**DOI:** 10.64898/2026.03.02.26347127

**Authors:** Min Shi, Leila Gheisi, Hao Zheng, Raju Gottumukkala, Nussdorf Jonathan, Grayson W. Armstrong, Lucy Q. Shen, Mengyu Wang

## Abstract

Early screening for glaucoma and diabetic retinopathy (DR) is critical to prevent irreversible vision loss, yet remains inaccessible to many underserved populations. However, AI models trained on hospital-grade fundus images often generalize poorly to low-cost images acquired with portable devices such as smartphones. We proposed CausalFund, a causally inspired learning framework for training AI models that enable reliable low resource screening from easily acquired non-clinical images. CausalFund disentangles disease-relevant retinal features from spurious image factors to achieve domain generalizable screening across clinical and nonclinical settings. We integrated CausalFund with seven deep learning backbones for glaucoma and DR screening from portable-device fundus images, including lightweight architectures suitable for on-device deployment. Across diverse experimental settings and image quality conditions, CausalFund improved smartphone-domain AUC for most backbones while maintaining comparable hospital-domain performance, and achieved a more balanced sensitivity-specificity trade-off than baselines. As a model-agnostic framework, CausalFund could be extended to other diseases and low-resource settings characterized by degraded or non-standard imaging.

## Introduction

Glaucoma and diabetic retinopathy (DR) are leading causes of preventable vision impairment worldwide,^1,2^ and both diseases benefit substantially from early detection and timely referral or treatment.^3–5^ However, comprehensive eye exams and high quality retinal imaging infrastructure are often concentrated in well-resourced clinical centers, leaving financially disadvantaged families and individuals in rural areas without routine examinations.^6,7^ This access gap motivates reliable and low cost solutions that can function outside traditional ophthalmology clinics.

In recent years, artificial intelligence (AI)-based approaches are widely utilized to automatically detect glaucoma and DR using retinal imaging such as optical coherence tomography (OCT) and fundus images (**Figure 1a**), which hold significant promise to alleviate clinical resource burdens and enable affordable screening.^8–11^ However, existing AI models depend heavily on high quality retinal images captured from costly imaging devices,^12,13^ such as those from Topcon and Zeiss, typically available in hospital and ophthalmology clinic-based settings. Such domain-dependent models trained on curated clinical datasets may fail to transfer to new acquisition conditions, devices, and populations.^14,15^ In fundus imaging, the shift from hospital-grade cameras to portable devices (e.g., smartphones) may introduce changes in field of view, illumination, sharpness, color response, and imaging artifacts,^16,17^ which can substantially degrade model reliability at deployment for cost effective screening. As a result, current AI screening models may not be easily deployed in low resource environments where hospital grade fundus cameras are unavailable. This significantly limits the reach of AI screening solutions to broader populations. An effective way to reduce this gap is to collect labeled target domain data and retrain or finetune models, but this is often impractical in low resource settings where labeled data and consistent acquisition protocols are difficult to obtain. Moreover, conventional training can encourage reliance on spurious factors, such as patient demographics, device signatures, compression artifacts, or quality related cues, that correlate with labels in the training environment but are unstable across domains.^18–21^ This is especially problematic for nonclinical fundus screening, where capture conditions are uncontrolled and image quality can vary substantially.

**Figure 1:**
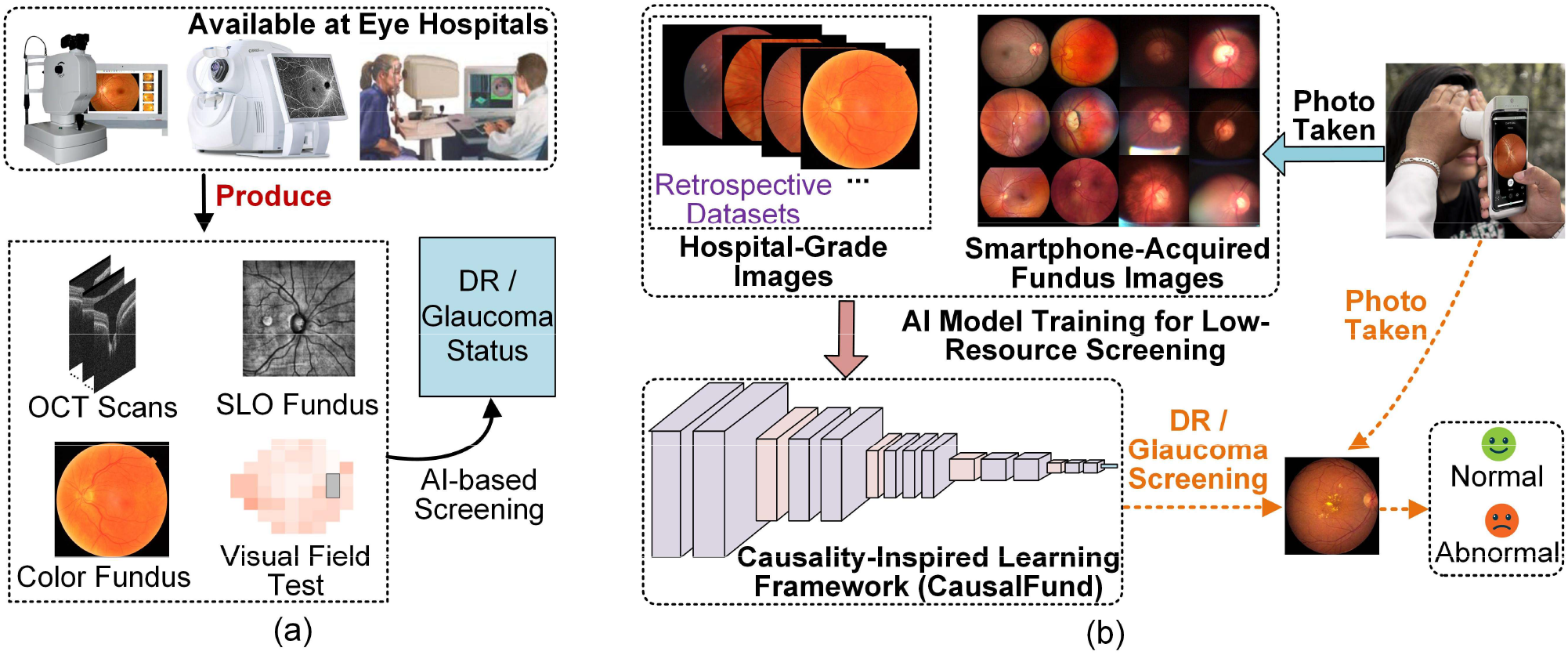
Illustration of low-resource screening. (a) Existing AI-based screening using hospital-based imaging devices. (b) Cost-effective AI for low-resource screening.

This work aims to develop robust fundus screening models that generalize from hospital grade clinical images to nonclinical, low-cost images acquired with smartphones or portable imaging devices (**Figure 1b**). We argue that a key ingredient is to explicitly discourage models from encoding domain specific nuisance variation, and instead promote representations aligned with disease relevant retinal structure,^22^ such as the cup-to-disc ratio and neuroretinal rim thinning.

Our perspective is inspired by broader insights from causal machine learning,^23–26^ which suggest that relationships between causal features and labels remain stable across environments, and that learning invariant predictors can substantially improve robustness when spurious correlations shift across domains. Accordingly, we proposed a causality-inspired framework called CausalFund to train cost-effective AI models for low-resource glaucoma and DR screening. CausalFund pairs each latent representation with a perturbed counterpart (**Figure 2**) and encourages prediction consistency between them, thereby discouraging reliance on unstable, domain-dependent image factors and improving generalization from hospital-grade retinal fundus images to portable device images. We integrated CausalFund with seven deep learning backbones and evaluated it for both glaucoma and DR screening under multiple experimental settings and image quality conditions. Across different experimental settings, CausalFund improved smartphone-domain screening performance and yielded a more balanced sensitivity-specificity trade off than conventional AI models, with gains maintained across mild, moderate, and severe image quality degradations.

**Figure 2:**
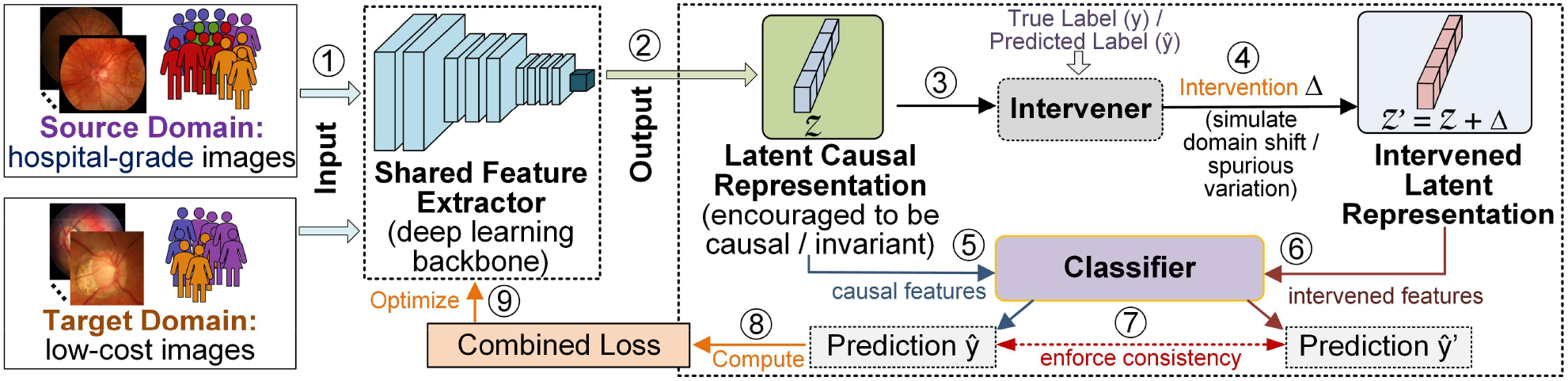
The proposed CausalFund framework. A shared backbone maps fundus images to a latent representation Z. A label-conditioned intervener adds a perturbation Δ to form Z′. A shared classifier predicts from Z and Z′. Consistency between predictions and the combined loss encourage stable and label-preserving representations. At inference, only the prediction from Z is used.

## Results

### Dataset Collection

We curated two domain-paired datasets for evaluating domain generalization in retinal fundus screening, including one for glaucoma and another for DR screening. For each task, we paired a hospital-based fundus dataset (e.g., higher-quality camera acquisitions) with a portable or smartphone dataset (e.g., lower-cost acquisitions with greater variability in illumination, focus, and field of view). All datasets were obtained from publicly available sources and used in accordance with their respective licenses and data-use policies. For the glaucoma dataset in hospital domain, we used the Multichannel Glaucoma Benchmark Dataset, which provides fundus images and dataset annotations for glaucoma classification. For the smartphone domain, we used the Brazil Glaucoma (BrG) dataset, which contains retinal images acquired with portable or smartphone based devices and corresponding glaucoma labels. After preprocessing and patient-level splitting, the hospital glaucoma domain contained 12,316 images and the smartphone glaucoma domain contained 2,000 images (**Table 1**). We treated both datasets as a binary classification task (healthy vs glaucoma) using the dataset-provided annotations.

**Table 1.**
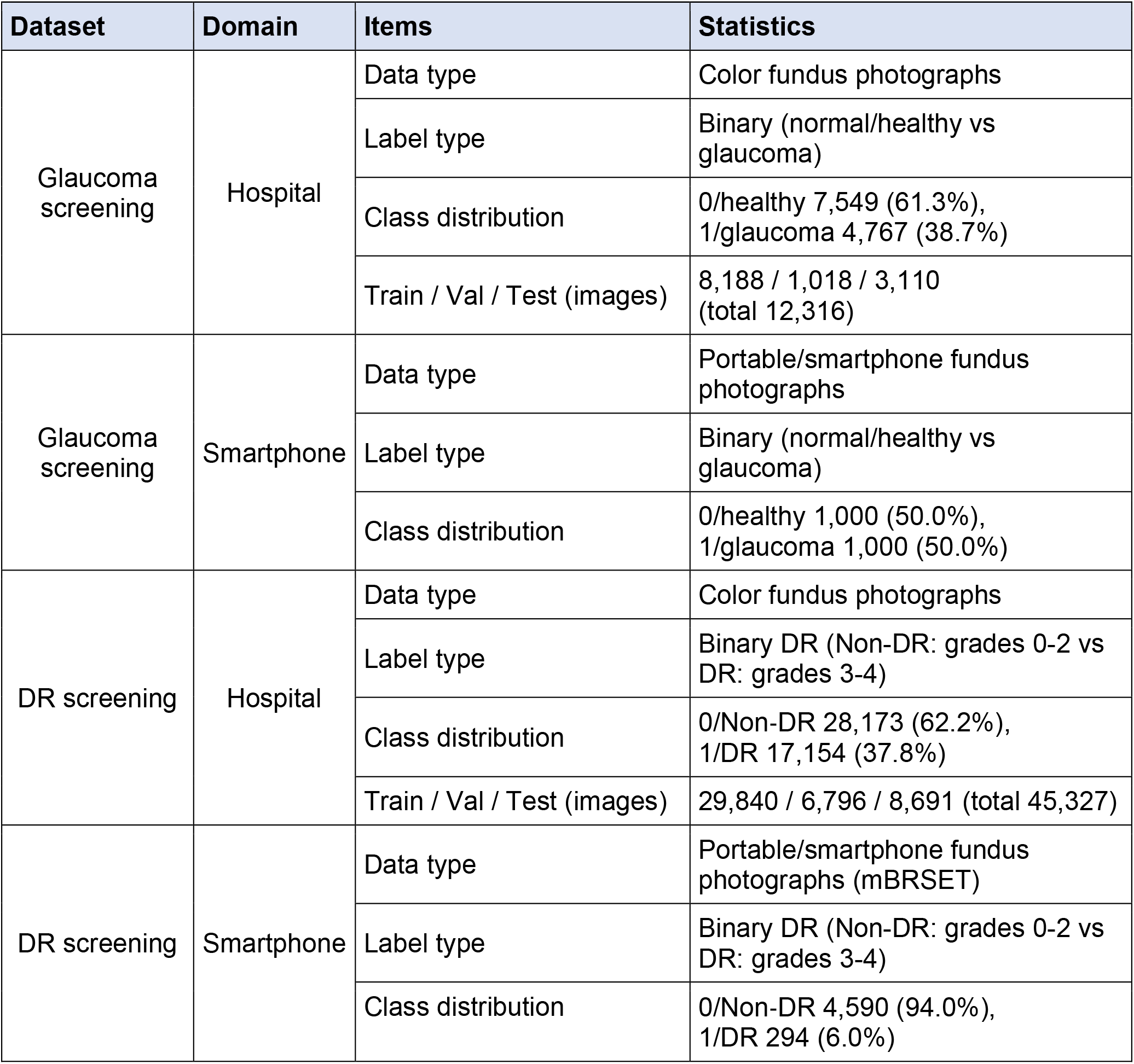
Dataset characteristics.

For the DR dataset in the hospital domain, we used a combined fundus dataset aggregating multiple widely used hospital-grade DR sources (APTOS, DDR, IDRiD, EyePACS, and Messidor). For the smartphone domain, we used the Mobile Brazilian Retinal Dataset (mBRSET) hosted on PhysioNet, which comprises retinal fundus photographs acquired using a handheld portable camera integrated with a smartphone platform and labeled for DR severity using standard DR grading. DR grades follow the conventional five-level scale (0-4: no DR, mild NPDR, moderate NPDR, severe NPDR, proliferative DR). After preprocessing and splitting, the hospital DR domain contained 45,327 images, and the smartphone DR domain contained 4,884 images (**Table 1**).

### Glaucoma Screening Results

We compared CausalFund with conventional empirical risk minimization (ERM), defined as standard supervised cross-entropy training of the same backbone classifier without CausalFund intervention losses, based on seven backbone architectures, namely five standard models (ResNet, DenseNet, VGG, EfficientNet, and ViT) and two lightweight models (MobileNet and SqueezeNet). CausalFund achieved higher smartphone domain AUC than ERM for six of the seven backbones (**Figure 3a-d**). In the hospital domain, ERM AUC ranged from 0.847 to 0.873, whereas CausalFund ranged from 0.850 to 0.862. For example, VGG changed from 0.851 to 0.859 (p = 0.103) and DenseNet from 0.860 to 0.862 (p = 0.679), whereas EfficientNet (0.872 to 0.859, p = 0.012), ResNet (0.865 to 0.855, p < 0.01), and ViT (0.873 to 0.861, p < 0.001) showed small decreases. In the smartphone domain, ERM AUC ranged from 0.726 to 0.820, while CausalFund ranged from 0.596 to 0.844, including significant improvements of ResNet from 0.759 to 0.844 (p < 0.001), MobileNet from 0.743 to 0.797 (p < 0.001), and VGG from 0.767 to 0.799 (p < 0.01).

**Figure 3:**
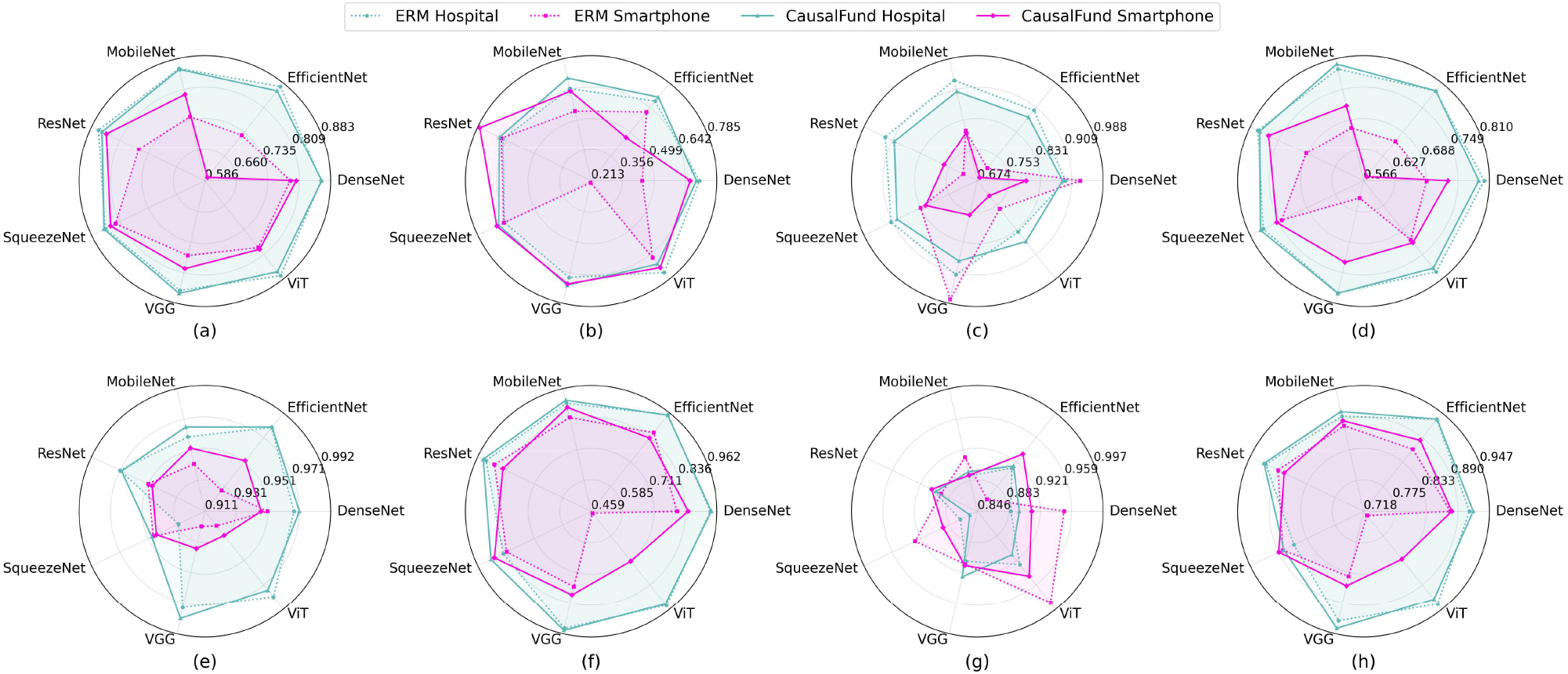
Glaucoma and DR screening results of ERM and CausalFund combined with various backbones, validated on hospital and smartphone-based fundus images. (a) Glaucoma screening AUC. (b) Glaucoma screening Sensitivity. (c) Glaucoma screening Specificity. (d) Sensitivity/specificity averaged results of glaucoma screening. (e) DR screening AUC. (f) DR screening Sensitivity. (g) DR screening Specificity. (h) Sensitivity/specificity averaged results of DR screening.

For sensitivity, hospital domain ERM values ranged from 0.646 to 0.747, compared with 0.676 to 0.703 for CausalFund (**Figure 3b**). Smartphone domain sensitivity under ERM ranged from 0.223 to 0.666, while CausalFund ranged from 0.468 to 0.775. For specificity, hospital domain ERM ranged from 0.838 to 0.933 and CausalFund ranged from 0.868 to 0.904 (**Figure 3c**). Smartphone-domain specificity ranged from 0.713 to 0.978 for ERM and from 0.684 to 0.817 for CausalFund. CausalFund thus reduced the sensitivity-specificity imbalance of ERM on smartphone images. For example, VGG under ERM reached a specificity of 0.978 but a sensitivity of only 0.223, compared with 0.762 and 0.695 under CausalFund. When averaging sensitivity and specificity (**Figure 3d**), for hospital-domain standard models, ERM averages ranged from 0.789 to 0.800 (e.g., 0.794 for ResNet, 0.792 for ViT), compared with 0.783 to 0.791 for CausalFund (e.g., 0.790 for ResNet, 0.783 for ViT). For hospital domain lightweight models, ERM achieved 0.790 for MobileNet and 0.782 for SqueezeNet, compared with 0.800 and 0.788 for CausalFund. In the smartphone domain, ERM averages for standard backbones ranged from 0.601 to 0.713 (e.g., 0.601 for VGG, 0.690 for ResNet), while CausalFund ranged from 0.576 to 0.770 (e.g., 0.728 for VGG, 0.770 for ResNet). For lightweight models, ERM averaged 0.673 for MobileNet and 0.742 for SqueezeNet, compared with 0.717 and 0.753 for CausalFund.

### Diabetic Retinopathy Screening Results

In the hospital domain, ERM AUC ranged from 0.930 to 0.981, whereas CausalFund ranged from 0.949 to 0.982 (**Figure 3e**). For example, SqueezeNet improved from 0.930 to 0.949 (p < 0.001), VGG from 0.974 to 0.982 (p < 0.001), and DenseNet from 0.968 to 0.972 (p < 0.01), EfficientNet was unchanged, and ViT decreased slightly from 0.981 to 0.976 (p < 0.001). In the smartphone domain, ERM AUC ranged from 0.921 to 0.951, while CausalFund ranged from 0.931 to 0.953 (**Figure 3e**), including improvements for EfficientNet from 0.928 to 0.952 (p < 0.001), MobileNet from 0.942 to 0.953 (p = 0.010), and VGG from 0.921 to 0.936 (p = 0.025), whereas DenseNet, ResNet, and SqueezeNet did not differ significantly between methods (p > 0.4).

For sensitivity (**Figure 3f**), hospital-domain ERM values ranged from 0.849 to 0.952, compared with 0.903 to 0.950 for CausalFund. Smartphone-domain sensitivity ranged from 0.469 to 0.888 under ERM and from 0.714 to 0.888 under CausalFund. For specificity (**Figure 3g**), hospital-domain ERM ranged from 0.868 to 0.928, while CausalFund ranged from 0.856 to 0.926. Smartphone-domain specificity ranged from 0.864 to 0.987 for ERM and from 0.890 to 0.946 for CausalFund. When averaging sensitivity and specificity (**Figure 3h**), hospital domain ERM values ranged from 0.859 to 0.935, while CausalFund ranged from 0.879 to 0.937. In the smartphone domain, ERM averages ranged from 0.728 to 0.891, compared with 0.830 to 0.889 for CausalFund. The largest gain was for ViT (0.728 to 0.830, p < 0.001), for which ERM reached a specificity of 0.987 but a sensitivity of only 0.469, compared with 0.946 and 0.714 under

CausalFund. For lightweight models specifically, hospital-domain ERM achieved 0.896 for MobileNet and 0.859 for SqueezeNet, compared with 0.904 and 0.879 for CausalFund. In the smartphone domain, ERM averaged 0.878 for MobileNet and 0.881 for SqueezeNet, compared with 0.887 and 0.889 for CausalFund.

### Impact of Smartphone Image Quality Degradation on Screening Results

Glaucoma screening results showed that the gains from CausalFund over ERM under image-quality degradation were concentrated in smartphone-domain screening (**Figure 4**). Mean improvements in smartphone domain AUC across all seven backbones were positive at all three degradation levels (mild 0.0622, moderate 0.0362, severe 0.0388; p < 0.001), hospital domain

**Figure 4:**
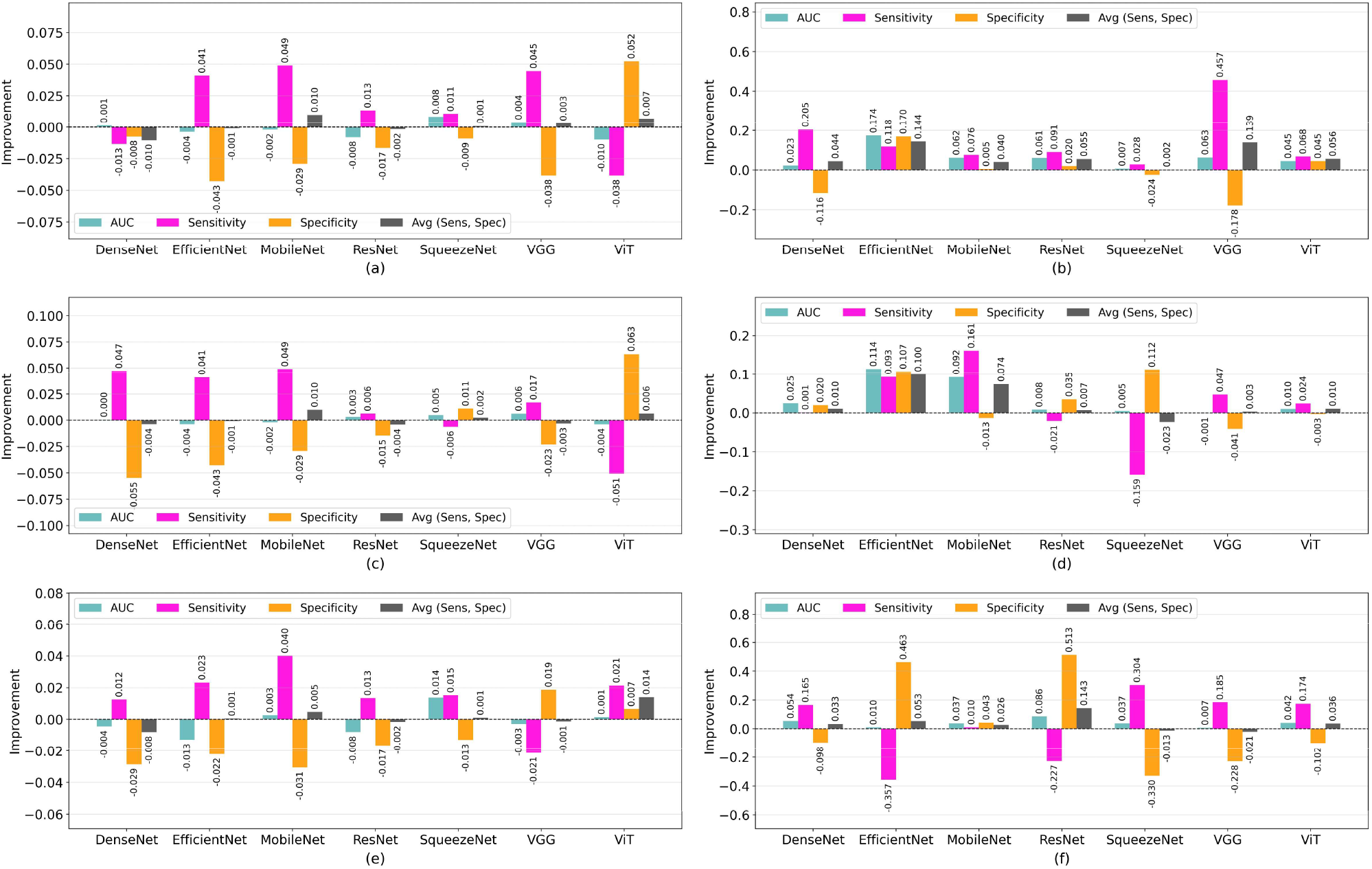
Glaucoma screening improvements of CausalFund over ERM after applying different levels (mild, moderate and severe) of degradation on smartphone images. (a) Hospital screening improvements after mild degradation. (b) Smartphone screening improvements after mild degradation. (c) Hospital screening improvements after moderate degradation. (d) Smartphone screening improvements after moderate degradation. (e) Hospital screening improvements after severe degradation. (f) Smartphone screening improvements after severe degradation. The improvement was computed by CausalFund - ERM.

AUC differences were small and not statistically significant (p > 0.29). Improvements of averaged sensitivity-specificity followed the same pattern, with smartphone domain gains of 0.0689 (mild), 0.0259 (moderate), and 0.0368 (severe) (p < 0.001) and near zero hospital domain changes (0.0010 to 0.0013; p > 0.5). On smartphone images, sensitivity gains were largest under mild degradation (0.1490), with smaller gains under moderate (0.0209) and severe (0.0363) degradation (p < 0.001), while specificity improved under moderate (0.0310) and severe (0.0373) degradation (p < 0.001) and decreased slightly under mild degradation (−0.0111, p = 0.045). In the hospital domain, CausalFund yielded slightly higher sensitivity (0.0148 to 0.0153) and slightly lower specificity (−0.0130 to −0.0123; p < 0.001). Overall, the glaucoma screening results indicate that CausalFund improved smartphone domain screening at every degradation level while hospital-domain AUC differences remained small and not statistically significant.

For DR, the gains from CausalFund were likewise concentrated in smartphone domain screening (**Figure 5**). Mean smartphone domain AUC improvements across all seven backbones were positive at all three degradation levels and increased with severity (mild 0.0052, p < 0.01; moderate 0.0098 and severe 0.0254, p < 0.001), whereas hospital domain AUC was unchanged under mild degradation and slightly lower under moderate and severe degradation (p < 0.001). The averaged sensitivity-specificity metric improved in the smartphone domain at all three levels (mild 0.0185, moderate 0.0418, severe 0.0271; p < 0.001), while hospital-domain changes were small and negative (−0.0011 to −0.0078). On smartphone images, sensitivity gains were largest under moderate degradation (0.0831, p < 0.001) and specificity gains were largest under severe degradation (0.0809, p < 0.001), where sensitivity decreased (−0.0267, p < 0.01). These results showed that CausalFund improved DR screening primarily in the smartphone domain under image-quality degradation.

**Figure 5:**
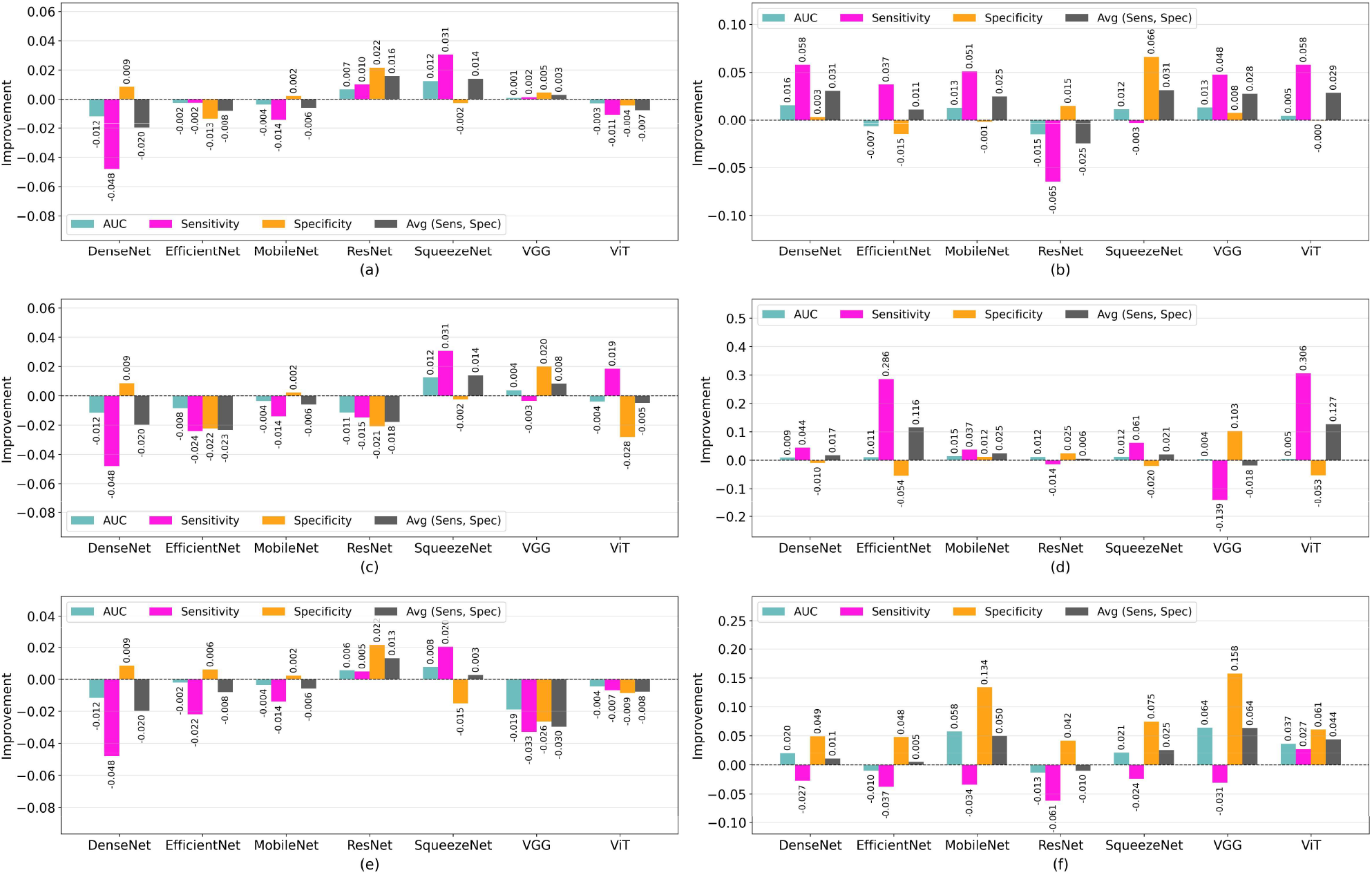
DR screening improvements of CausalFund over ERM after applying different levels (mild, moderate and severe) of degradation on smartphone images. (a) Hospital screening improvements after mild degradation. (b) Smartphone screening improvements after mild degradation. (c) Hospital screening improvements after moderate degradation. (d) Smartphone screening improvements after moderate degradation. (e) Hospital screening improvements after severe degradation. (f) Smartphone screening improvements after severe degradation. The improvement was computed by CausalFund - ERM.

Across all seven backbones, CausalFund maintained a higher mean smartphone domain AUC than ERM at every degradation level for both glaucoma (mild 0.835 vs. 0.772, moderate 0.849 vs. 0.813, severe 0.714 vs. 0.676) and DR (mild 0.947 vs. 0.941, moderate 0.938 vs. 0.928, severe 0.870 vs. 0.844) (**Figure 6a-f**). From mild to severe degradation (**Figure 6g-h**), the mean DR AUC declined more slowly with CausalFund than with ERM (linear-fit slope −0.039 vs. −0.049 per level), whereas for glaucoma CausalFund declined somewhat faster (−0.060 vs. −0.048 per level) while remaining above ERM at every level. As an example, from mild to severe smartphone image quality variations (**Figure 7a**), the baseline ERM approach learned from varied regions caused by changing image qualities (**Figure 7b**), whereas CausalFund consistently focused on learning features at the optic nerve head area for glaucoma detection (**Figure 7c**).

**Figure 6:**
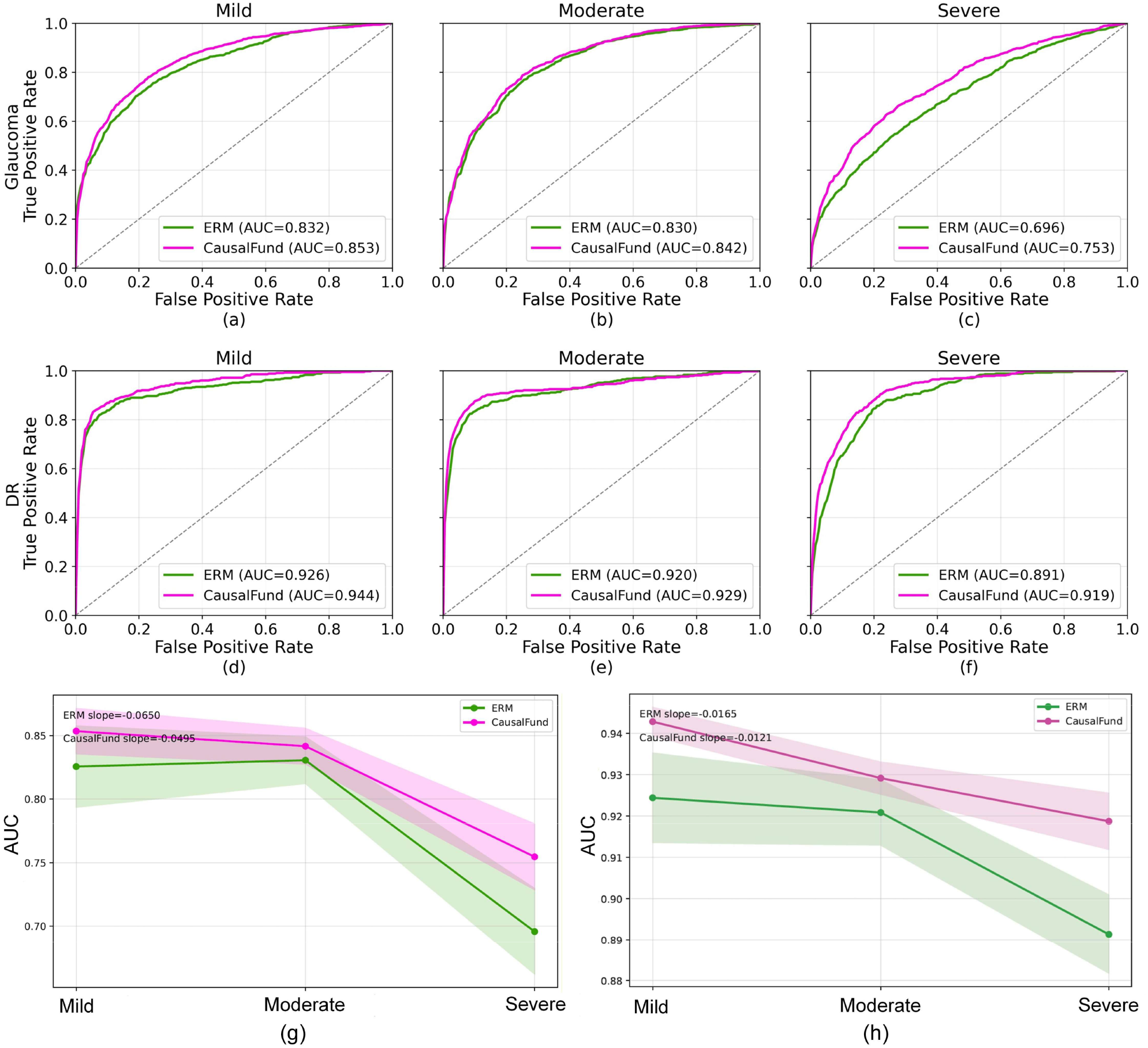
Smartphone ROC curves under progressive image-quality degradation for glaucoma (top) and DR (bottom). Columns correspond to mild, moderate, and severe degradation. Curves are mean ROCs across the seven backbones. (a-c) Glaucoma. (d-f) DR. (g) Mean AUC results of glaucoma screening. (h) Mean AUC results of DR screening. Slopes are linear fits to the three quality levels and shaded bands indicating standard error of the mean across all backbones for ERM and CausalFund at mild, moderate, and severe degradation levels.

**Figure 7:**
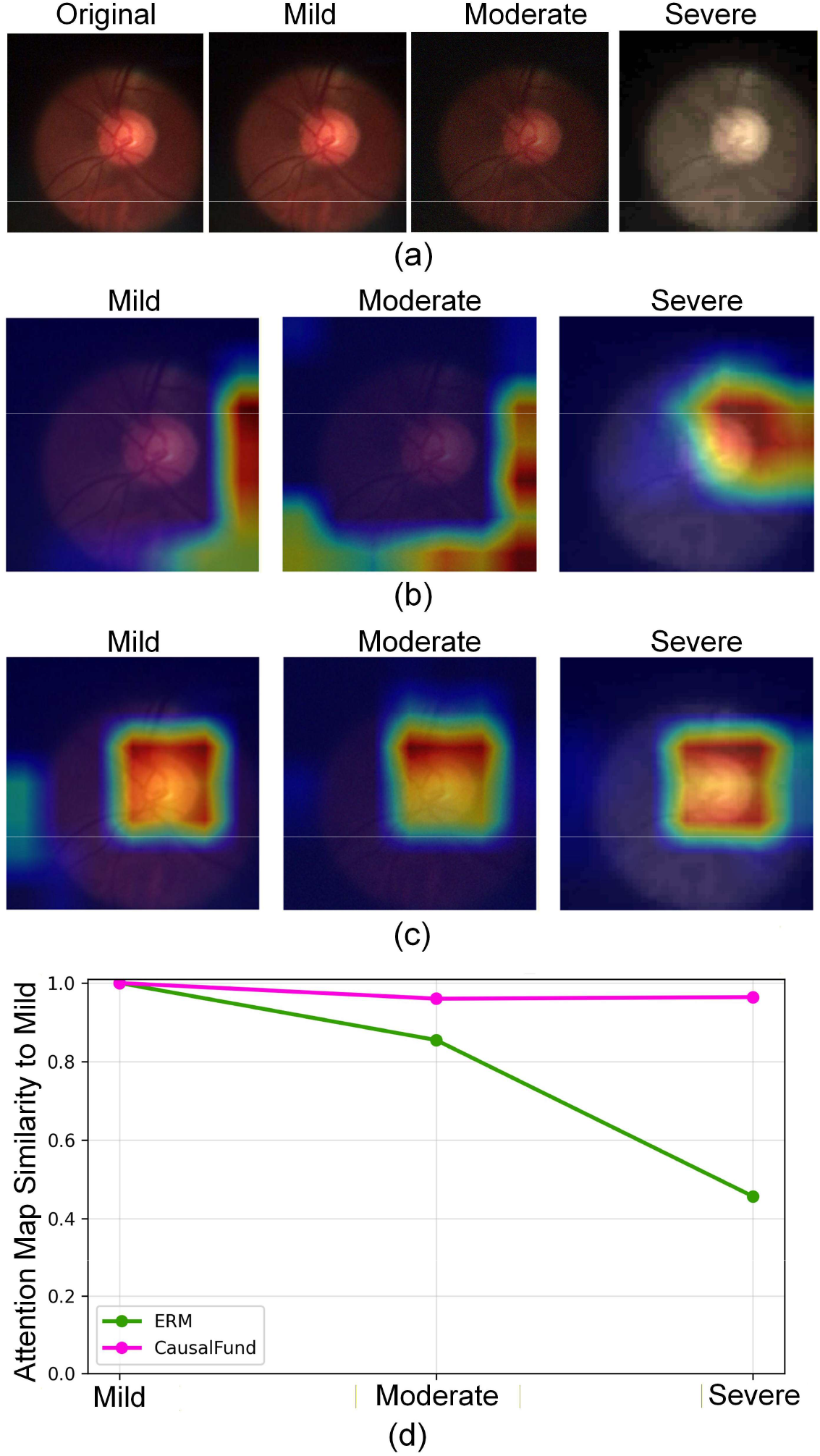
Grad-CAM comparison between ERM and CausalFund in smartphone-based glaucoma screening, and respective attention stabilities under progressive image-quality degradation - mild, moderate and severe. (a) The original smartphone image and images after applying mild, moderate and severe image-quality degradations. (b) Grad-CAM activation map of ERM (MobileNet backbone) under mild, moderate, and severe smartphone-quality degradations. Warmer colors indicate stronger Grad-CAM activation. (c) Grad-CAM activation map of CausalFund. CausalFund generally preserves attention on disease-relevant retinal regions more consistently as image quality deteriorates. (d) Cosine similarity was computed between the Grad-CAM at each quality level and the corresponding mild-quality Grad-CAM (reference).

### Ablation Study

We assessed the contribution of the CausalFund components. The ablation used the reported EfficientNet-B0 configuration for DR screening (**Figure 8**). Full CausalFund achieved the highest smartphone AUC (0.952). Removing consistency regularization reduced AUC to 0.946 (P = 0.161). Replacing the class conditional prior, using classification loss alone, replacing the learned perturbation with a random control, or removing the KL term reduced AUC to 0.932, 0.928, 0.925, and 0.920, respectively (all P < 0.001). The standard prior increased sensitivity (0.908 vs 0.847) while reducing specificity (0.833 vs 0.918). Full CausalFund retained the highest specificity (0.918).

**Figure 8:**
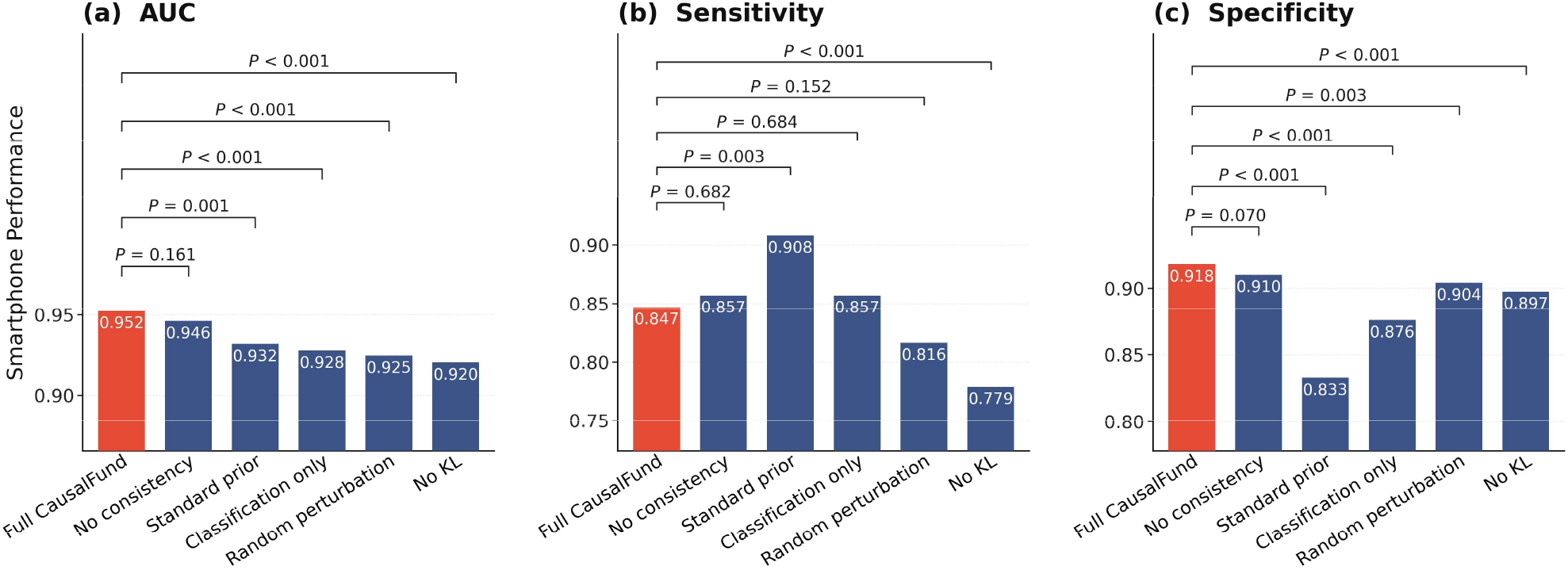
Component ablation of CausalFund. (a) AUC. (b) Sensitivity. (c) Specificity. Brackets show two-sided P values versus Full CausalFund.

## Discussion

Recent years have witnessed a surge of AI models for automatically detecting retinal diseases such as glaucoma and DR.^27–29^ Current works consistently demonstrate that AI-based screening has considerable potential to reduce clinical care burden and cost to benefit broader populations, especially those from remote and low-resource areas.^7,30^ However, the adoption of AI in real-world clinical environments remains hesitated and lagged behind. An undeniable factor lies in the fact that most state-of-the-art AI models are trained and evaluated using hospital grade data from quality-guaranteed acquisition conditions, which generally suffer from poor generalizability to low resource settings where acquisition conditions and qualities are uncontrollable.^24,31^ This study aims to mitigate this gap by training generalizable AI models for glaucoma and DR using low cost fundus images acquired from portable devices and smartphones. The proposed CausalFund framework (**Figure 2**) aims to train domain-generalizable AI models and represents an effort to translate AI advances into realistic applications for cost-effective screening.

Our study showed that models trained in hospital domain suffer from significant performance degradation when applied in smartphone domain, which was especially prominent in poor imaging conditions. For example, under severe smartphone image quality degradation, the averaged glaucoma AUC gap between hospital and smartphone domains based on ERM across all seven backbones was 0.184 (p < 0.001), while this gap was significantly reduced to 0.143 (p < 0.001) with CausalFund, which encouraged predictions to remain stable under latent perturbations. For DR, the corresponding gap decreased from 0.123 to 0.094 (p < 0.001). For both glaucoma and DR screening, CausalFund maintained higher smartphone domain AUC than ERM at every degradation level (**Figure 6**), its degradation trend was flatter than that of ERM for DR, indicating improved robustness to image-quality shifts. We attribute this advantage to the perturbation-consistency objective, which discourages reliance on unstable image factors. Disease relevant structures such as the cup-to-disc ratio and neuroretinal rim are expected to remain more stable across acquisition conditions. The case study showed that CausalFund was more robust to varying imaging conditions (e.g., spurious factors) and demonstrated stable learning capacity across domains (**Figure 7**). For example, the Grad-CAM analysis is consistent with greater attention stability around the optic nerve head. This is critical for low-resource settings where imaging conditions are often unpredictable and a generalizable model is desired.

Despite the advantages, our study has several limitations. First, domain generalization seeks to improve performance under distribution shifts without updating model parameters on the deployment domain.^32^ Many established approaches, including invariant risk minimization,^33^ GroupDRO,^34^ and domain-alignment methods,^35^ are formulated using multiple labeled source environments. Our study instead uses single-source training in that one hospital domain supplies all parameter updates, while the smartphone domain is held out from model training. Methods designed specifically for single source domain generalization exists but were not evaluated here. Accordingly, our experiments assess CausalFund relative to matched ERM models across architectures and do not establish state-of-the-art performance among domain generalization methods. Second, image quality shifts were modeled using predefined mild, moderate, severe augmentation pipelines. Although useful for controlled analysis, these synthetic degradations cannot capture all artifacts seen in practice (e.g., mixed blur, illumination instability, compression, and operator-dependent errors). Third, we focused on two tasks, glaucoma and DR, and a fixed set of backbones, so generalizability to other ophthalmic conditions, model families, and institutions remains to be established. Fourth, CausalFund is a causality-inspired robustness framework rather than a method for identifying causal representations. Its latent perturbation and consistency losses encourage prediction stability but do not constitute observed interventions on acquisition variables, guarantee invariance to specific nuisance factors, or demonstrate that individual latent variables correspond to causal retinal mechanisms. The mild, moderate, and severe degradations are controlled tests under simulated acquisition shifts, and the Grad-CAM and attention-stability analyses provide supportive rather than definitive causal evidence. Fifth, although separating the clean prediction pathway from class conditioning and applying shared-classifier consistency constraints reduce direct label leakage, they do not mathematically exclude class-conditional shortcut learning during optimization. Comparing class-conditioned and unconditioned intervention modules is an important direction for future ablation studies. Finally, we did not perform prospective clinical validation, fairness audits across demographic subgroups, and deployment level analyses of latency and integration, which are important next steps before real-world adoption.

In conclusion, current deep learning models often use images from hospital-based devices, which can demonstrate poor performance in low-resource settings due to uncontrolled, degraded image qualities. This work proposed a causality-inspired framework to encourage deep learning models to rely on disease relevant retinal features that remain stable across fundus imaging domains for robust screening performance. Empirical results and comparisons with traditional approaches demonstrated the effectiveness of CausalFund. The framework could be extended to other diseases to potentially achieve robust screening in low resource imaging settings.

## Methods

Four datasets were used to develop and evaluate deep learning models for glaucoma and DR screening. This study was reviewed and determined to be exempt by the Institutional Review Board at University of Louisiana ay Lafayette (IRB No. IRB-25-088-CMPS-OLVR). The research involved secondary analysis of publicly available data. The requirement for informed consent was waived. The study was conducted in accordance with the Declaration of Helsinki.

### Dataset Description

For glaucoma, we used the dataset-provided annotations and formulated the task as binary classification (normal vs. glaucoma). We enforced patient-level splitting to prevent identity leakage. For the hospital glaucoma dataset, we parsed the provided metadata to derive patient identifiers and labels, removed mixed-label patients (i.e., patients with conflicting labels across their images), and then performed a patient-level split (70%, 10%, 20%) into train/validation/test. For DR, labels follow the standard five-level grading scale (0-4: no DR, mild NPDR, moderate NPDR, severe NPDR, proliferative DR). We binarized DR severity into Non-DR (grades 0-2) versus DR (grades 3-4) and enforced this mapping consistently across the hospital and smartphone DR datasets. All images were loaded as RGB and resized to the model input size during training, and standard label-preserving training augmentations (random crop, horizontal flip, ±15° rotation, and color jitter) were applied to the training split only for both ERM and CausalFund. For both glaucoma and DR, all smartphone images were used only for testing.

### The Proposed CausalFund Framework

CausalFund is a causality inspired domain generalization framework designed to improve robustness when deploying fundus image screening models across acquisition domains, such as from hospital-grade cameras to smartphone devices in this study. The main idea is to discourage reliance on domain-specific factors by explicitly training the model to be stable to controlled perturbations of its internal representation that simulate such spurious variation. CausalFund is motivated by the hypothesis that disease relevant retinal structure is more stable across acquisition environments than nuisance image characteristics such as illumination, contrast, blur, sensor noise, and compression. The learned latent perturbation is intended to expose the classifier to variation in its internal representation, while the consistency objective penalizes changes in prediction under that variation. Because the perturbation is not supervised by measured acquisition variables, this objective encourages robustness to potential nuisance information but does not identify causal factors or guarantee that individual latent variables correspond to causal retinal mechanisms. Given an input fundus image, CausalFund contains the following learning and optimization processes (**Figure 2**). First, the shared feature extractor, implemented as a deep learning backbone, learns image features into a latent representation *Z*. Then, an intervener, implemented as a three-layer multilayer perceptron (MLP) network, generates a label-conditioned (e.g., conditioned on the true label *y* or model-predicted label *ŷ* of the image) perturbation Δ intended to mimic domain shift in spurious factors while preserving the class semantics. This will result in an intervened latent representation *Z*^′^ = *Z* + Δ. Using the label (or predicted label) as the condition encourages the intervener to change nuisance characteristics without crossing the decision boundary into another disease class. Then, a shared classifier aims to predict class labels *y* and *y*^′^ from latent representation *Z* and intervened representation, respectively. The model enforces predicted class labels *ŷ* and *ŷ*^′^ to be consistent, by which the model is optimized to use stable disease-relevant representation *Z* for prediction without being affected by spurious features controlled by Δ. Taken together, CausalFund optimizes a combined loss that encourages both predictive performance and invariance to interventions:

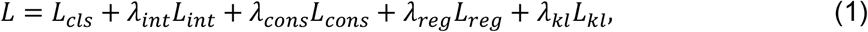

where *L*_*cls*_ = *CE*(*ŷ, y*) is the standard cross-entropy classification loss on the clean latent representation *Z. CE* represents the cross-entropy loss measuring how well the model predictions match the true class labels.^36^ *L*_*int*_ = *CE*(*ŷ*^′^, *y*) is the cross-entropy classification loss on the intervened latent *Z*^′^ = *Z* + Δ, encouraging the shared classifier to remain correct under intervention. 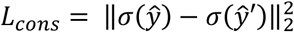 is a prediction consistency loss defined as the mean-squared error between sigmoid-transformed clean and intervened classifier logits, where *σ* is sigmoid activation function. 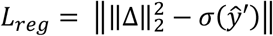 represents intervention regularization to constrain the magnitude of Δ with target scale *b* controlling typical intervention strength. *L*_*kl*_ = *D*_*kl*_(*z* || *p*(*z*|*y*)) represents latent regularization, i.e., the Kullback–Leibler divergence *D*_*kl*_, to discourage unstable latent representation learning, where *p*(*z*|*y*) is a class-conditional prior. *λ*_*int*_, *λ*_*cons*_, *λ*_*reg*_, and *λ*_*kl*_ are scalar weights. CausalFund conducts supervised training and optimizes with the Adam optimizer^37^. When the intervener is removed and only the classification loss is retained, Equation 1 reduces to the ERM objective.

#### Intervener architecture and training

Within CausalFund, a backbone featurizer first maps the image to a *d*-dimensional feature vector. Two linear heads then parameterize a latent Gaussian, and *Z* ∈ ℝ^*d*^ is sampled by the reparameterization trick (for ResNet-50, *d* = 2048). The intervener is a three-layer MLP on this latent code. It concatenates *Z* with a *C*-dimensional class-conditioning vector (*C* = number of disease classes) and passes this concatenated vector through hidden layers of width *d*/2 with ReLU activations to produce a residual Δ ∈ ℝ^*d*^ . The intervened latent is *Z*^′^ = *Z* + Δ. A shared linear classifier predicts disease labels from both *Z* and *Z*^′^. During supervised training on the hospital-domain training split, class conditioning uses the ground-truth one-hot label. Note that class conditioning does not enter the clean prediction pathway. The featurizer first computes *Z* from the image and the shared classifier produces the clean logits f(z) before the label is supplied to the intervener. The ground-truth label is used only by the auxiliary intervention branch and the class-conditional latent regularizer during training. The featurizer, latent heads, intervener, and classifier are optimized jointly with Adam under Equation 1. At inference, the reported screening score is the clean-branch output f(z), computed deterministically from the latent mean (no sampling). The intervener is not used to form the deployed prediction.

To evaluate CausalFund, we treated hospital and smartphone images as separate environments. In the primary domain generalization setting, models were trained on the hospital training split, while smartphone data were reserved for testing. The hospital domain was split at the patient level into 70%, 10%, and 20% for training, validation, and testing. Within each training run, the checkpoint was selected as the epoch with the highest hospital validation AUC. All reported test performance was computed from the selected checkpoint of each backbone. We evaluated CausalFund with both standard and lightweight deep learning backbones. Standard backbones included ResNet-50,^38^ DenseNet-121,^39^ EfficientNet-B0,^40^ VGG-16,^41^ and Vision Transformer (ViT-B/16),^42^ while lightweight backbones included MobileNet-V2^43^ and SqueezeNet-1.1.^44^

### Controlled Smartphone Image-Quality Degradation

To assess robustness under quality shift, we constructed three controlled degradation levels from the original smartphone images. Examples of the original smartphone images and the corresponding mild, moderate, and severe degradations are shown in Figure 9. The mild level applied Gaussian blur (radius = 1.2) and JPEG compression (quality = 50). The moderate level introduced downsample-upsample degradation (scale = 0.5), additive Gaussian noise (σ = 20), and reduced brightness (0.75) and contrast (0.85). The severe level combined motion blur (kernel size = 17), strong JPEG compression (quality = 15), and color distortion (hue shift = 0.07, saturation = 0.5). This progressively intensifying setup emulated realistic smartphone-specific artifacts, such as diminished sharpness, sensor noise, compression effects, and illumination or color instability. The mild, moderate, and severe degradations provide controlled robustness tests under simulated acquisition shifts. They alter resolution, blur, illumination and color, sensor noise, and compression while retaining the original disease labels. These transformations are not observed interventions on measured acquisition variables and are not used as evidence of causal identification.

**Figure 9:**
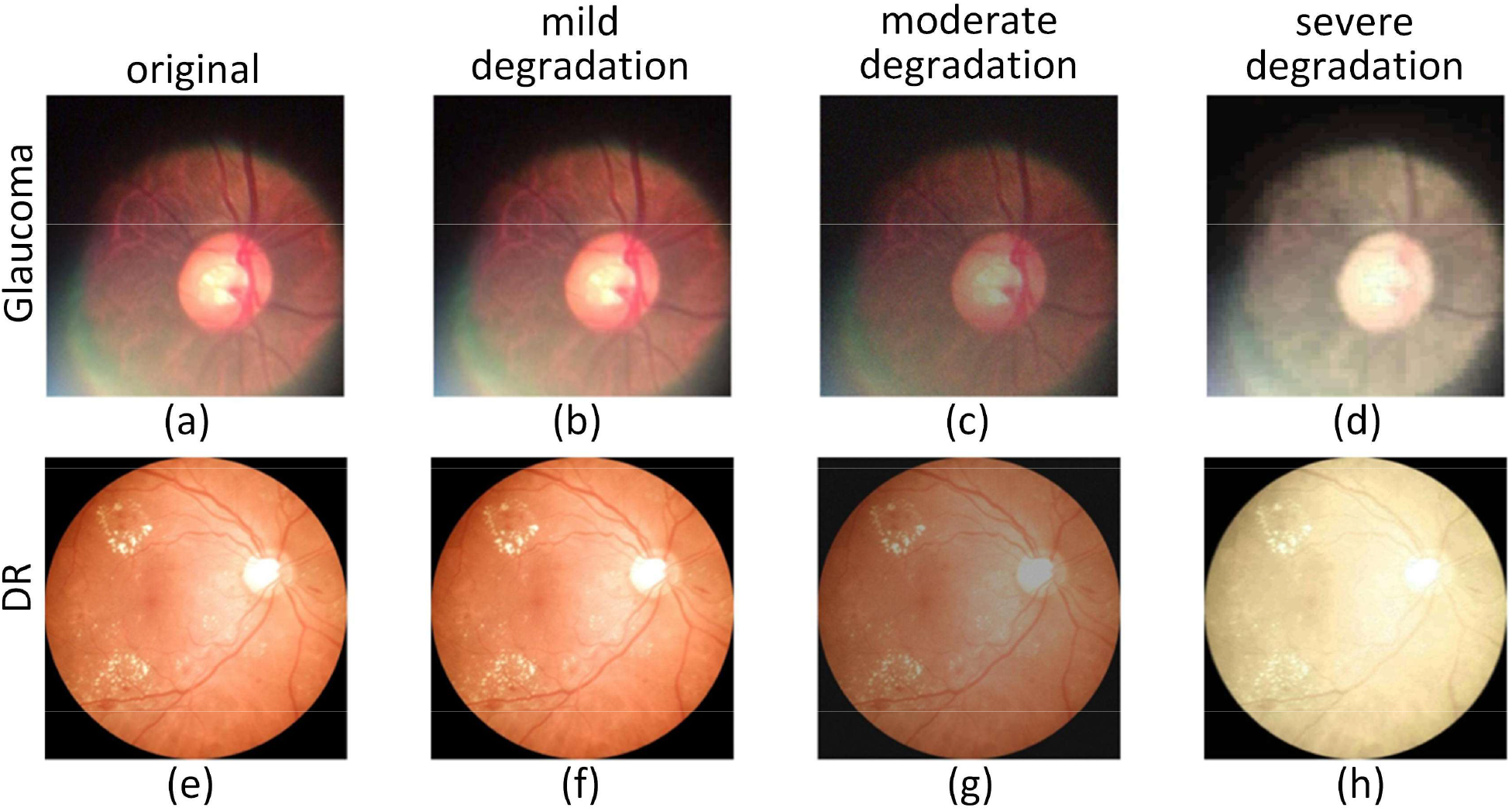
Example smartphone fundus images under controlled image-quality degradation. (a-d) Glaucoma original, mild, moderate, severe. (e-h) DR original, mild, moderate, severe.

### Comparative Methods, Evaluation Metrics and Statistical Analysis

We compared CausalFund against ERM baselines using a consistent training and evaluation protocol across both glaucoma and DR screening tasks. ERM denotes the standard supervised deep learning classification objective and does not introduce any additional statistical formulation beyond cross-entropy risk minimization. Formally, for a backbone feature extractor and classifier *f*_*θ*_ and hospital-domain training distribution 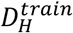, ERM minimizes:

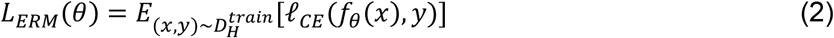

where ℓ_*CE*_ is the categorical cross-entropy loss. Equivalently, ERM retains only the classification term of CausalFund and omits the intervener network and all intervention-related losses in Equation 1. To isolate the contribution of CausalFund, ERM used identical backbone architectures, ImageNet-pretrained weights when applicable, image preprocessing, training-time data augmentation, optimizer (Adam), batch size, number of epochs, random seed, and validation-AUC checkpoint selection as CausalFund. The sole intentional difference was the training objective (Equation 2 versus Equation 1).

For each task, we evaluated the same seven backbones: ResNet, DenseNet, EfficientNet, VGG, ViT, MobileNet and SqueezeNet. Model performance was primarily assessed using the area under the receiver operating characteristic curve (AUC), sensitivity, and specificity. The threshold for sensitivity and specificity was calibrated independently in the hospital and smartphone domains to achieve the best balance of sensitivity and specificity. For each backbone and method, the best checkpoint was selected based on hospital-domain validation AUC, and all reported test-set metrics were computed using the selected checkpoint. For cross-domain evaluation, we reported results separately on hospital and smartphone test sets to quantify the domain generalization gap. Statistical analysis focused on comparing AUC and threshold-based metrics between CausalFund and ERM under matched backbones and experimental settings. We used paired comparisons across backbones and experimental repetitions. We estimated uncertainty via bootstrap resampling on the test set. Statistical significance was evaluated using two-sided tests with a significance threshold of p < 0.05.

### Parameter Settings

All models were trained using the same preprocessing and optimization pipeline to enable fair comparisons across methods and backbones. Standard data augmentation (random crop, horizontal flip, rotation, and color jitter) was applied to the training split only, while validation and test images were evaluated using deterministic preprocessing (resize and normalization) without augmentation. These image augmentations were applied identically to ERM and CausalFund and are distinct from the CausalFund intervener, which applies a learned residual perturbation in latent space after feature extraction rather than transforming the input image. The controlled mild, moderate, and severe smartphone degradations used in the robustness experiments are a separate evaluation protocol and were not used as the CausalFund intervention mechanism. We optimized model parameters using the Adam optimizer with weight decay disabled. Unless otherwise stated, models were trained for 40 epochs with a mini-batch size of 32 and a fixed random seed (seed = 0) to ensure reproducibility. CausalFund introduces additional parameters controlling the intervention module and regularization terms. We used the following default settings for CausalFund across tasks: learning rate 1 × 10^−5^, batch size 32, *b* = 2.0, *λ*_*int*_ = 0.5, *λ*_*cons*_ = 0.2, *λ*_*kl*_ = 0.01, and *λ*_*reg*_ = 0.2. These defaults were treated as a starting configuration rather than a single fixed setting for all models. For each backbone and task, we adjusted the intervention-related weights in a neighborhood of the defaults (primarily bias, λ_int, λ_consist, λ_reg, and λ_KL), guided by validation AUC and training stability, and then selected the final checkpoint by validation AUC.

## Data Availability

All datasets were from public sources and used under their respective licenses: hospital glaucoma (https://www.kaggle.com/datasets/deathtrooper/multichannel-glaucoma-benchmark-dataset), smartphone glaucoma (https://www.kaggle.com/datasets/clerimar/brasil-glaucoma-brg), hospital DR (https://www.kaggle.com/datasets/sehastrajits/fundus-aptosddridirdeyepacsmessidor), and smartphone DR (mBRSET, PhysioNet)

## Data availability

All datasets were from public sources and used under their respective licenses: hospital glaucoma (https://www.kaggle.com/datasets/deathtrooper/multichannel-glaucoma-benchmark-dataset), smartphone glaucoma (https://www.kaggle.com/datasets/clerimar/brasil-glaucoma-brg), hospital DR (https://www.kaggle.com/datasets/sehastrajits/fundus-aptosddridirdeyepacsmessidor), and smartphone DR (mBRSET, PhysioNet)^45^.

## Author Contributions

MS and MW, LQS conceived the study. MS and GL collected the data and developed deep learning models. MS and GL performed data processing and experiments. MS, GL, HZ, RG, and LQS performed results analysis. NJ, GWA and LQS contributed clinical expertise. MS supervised the work. All authors wrote and revised the manuscript. All authors have read and approved the manuscript.

## Competing Interests

The authors declare no competing interests.

## Notes

### Competing Interest Statement

The authors have declared no competing interest.

### Summary of Updates

New experiments added in the result sections; Discussion expanded to analyze the limitations of proposed approach; Figure 8 and Figure 9 were added; Method enriched with more details.

